# Chlorhexidine Bathing for Candida auris Decolonization among Adult Patients in Healthcare Settings: Protocol for a Systematic Review and Meta-Analysis

**DOI:** 10.1101/2024.12.20.24319367

**Authors:** Messaouda Belfakir, Moustafa Sherif, Balazs Adam

## Abstract

**Background:** Candida auris (C. auris) poses a significant threat in healthcare settings, characterized by its high morbidity and mortality rates. While the use of chlorhexidine bathing has been suggested as a potential strategy for C. auris decolonization in adult patients within healthcare settings, a comprehensive and systematic evaluation of its effectiveness and associated outcomes is notably lacking.

**Aim:** This study seeks to systematically assess the effectiveness of daily chlorhexidine bathing for Candida auris decolonization in adult patients within healthcare settings. The study’s primary objectives are to evaluate the impact of this intervention on reducing colonization rates, infection occurrences, and outbreak incidences, while concurrently evaluating any associated adverse events. The study’s secondary objectives are to identify adverse events, and to explore and quantify the effect sizes of potential risk factors, if identified, that may influence the outcomes of chlorhexidine bathing for C. auris decolonization.

**Methods and Analysis:** In adherence to the Preferred Reporting Items for Systematic Reviews and Meta-Analyses Protocol (PRISMA-P) guidelines, this protocol outlines the methodology for our systematic review and meta-analysis. The study commenced with an extensive presearch conducted from June to August 2023 on PubMed, followed by searches across other three key databases: Embase, Web of Science, and Scopus, in September 2023. The systematic search will encompass all available years of publication without applying any publication date filters. Records located in the literature search will be uploaded to the systematic review software Covidence to facilitate deduplication, blinded screening, and the selection of eligible studies. Two independent reviewers will rigorously screen records, extract data, and perform risk of bias assessments, with a third researcher resolving conflicts. The results will be synthesized narratively in summary tables, with the potential for meta-analysis contingent upon the findings, focusing on the effectiveness and adverse events of daily chlorhexidine bathing for C. auris decolonization in adult patients within healthcare settings. Additionally, we will investigate whether certain risk factors, if identified, have an impact on the outcomes by quantifying their effect sizes.

**Ethics and Dissemination:** The ethical framework of this systematic review obviates the need for ethics approval, as it relies exclusively on published research. The outcomes of this study will be disseminated via publication in a peer-reviewed journal, shared with stakeholders, and made publicly accessible.

**PROSPERO Registration:** CRD42023459048.

## Introduction

Candida auris (C. auris) represents a prominent pathogenic threat within healthcare environments, characterized by elevated rates of morbidity and mortality. This fungal pathogen, along with other fungal pathogens, contributes to approximately 13 million infections and 1.5 million deaths annually (1). A thorough examination of 4,733 cases reported across 33 countries revealed that the mortality rate associated with C. auris infection was 39%, with a higher rate of 45% for C. auris fungemia (2). Moreover, it is estimated that around 5% to 10% of individuals colonized with C. auris will eventually develop invasive disease (3,4).

C. auris initial discovery in 2009 marked the commencement of sporadic cases and widespread outbreaks worldwide. It presents a unique danger, particularly in the context of invasive infections, significantly impacting individuals in hospitals and nursing homes who contend with various underlying medical conditions. C. auris capacity to persist within patients for extended periods and withstand common healthcare disinfectants underscores its environmental resilience. Alarmingly, more than one-third of patients succumb within a month of being diagnosed with an invasive C. auris infection, as documented by the Centres for Disease Control and Prevention (5).

The precise factors contributing to the recent emergence of this fungus remain elusive. Genetic analyses suggest its simultaneous emergence across various continents, with the identification of five distinct C. auris clades in disparate geographical locations. Hospital-based C. auris infections and outbreaks have exhibited an upward trajectory in recent years, as observed in studies such as that conducted by Han, 2020 (6). Notably, the advent of the COVID-19 pandemic coincided with hospital outbreaks of multidrug-resistant organisms, further complicating the landscape for healthcare facilities (7). C. auris easily contaminates patient environments, persists, and causes major outbreaks. It resists cleaning due to dry biofilms. Vulnerable hospitalized patients, especially in ICUs, acquire it, often with fatal consequences (8).

C. auris can still be easily misidentified when using standard diagnosis methods. Definitive approaches for identifying C. auris encompass matrix-assisted laser desorption/ionization time-of-flight mass spectrometry (MALDI-TOF MS) and molecular techniques grounded in DNA sequencing. The management of C. auris within healthcare settings presents formidable challenges owing to several factors. Foremost among these is its resistance to multiple classes of antifungal agents, with a subset of isolates demonstrating resistance to all major antifungal medications. Specifically, 60-90% of C. auris strains manifest resistance to fluconazole, 10-30% exhibit elevated minimum inhibitory concentration values for amphotericin B, and up to 5% are considered resistant to echinocandins (9).

Furthermore, once C. auris established in patients, the fungus can endure in the environment and spread among individuals. Significantly, patient colonization by C. auris can persist over extended periods, even following treatment for invasive infections. Consequently, adherence to recommended infection control measures is imperative both during and after C. auris infection management. Given these formidable challenges, effective strategies for decolonization become paramount. However, data regarding the efficacy of such methods, including the utilization of chlorhexidine or topical antifungal agents, for patients with C. auris remain notably limited based on Centres for Disease Control and Prevention (CDC) (10).

Previous research, encompassing both in vitro and in vivo investigations, has indicated the efficacy of chlorhexidine gluconate (CHG) against Candida species, including C. auris. Interestingly, studies have shown that CHG is more effective against C. auris when combined with compounds like tea tree and lemongrass oil or antifungal drugs like fluconazole and terbinafine. On the other hand, using only 2% chlorhexidine resulted in less effective reduction of C. auris fungal burden compared to no treatment. However, after the utilization of the Advanced Performance Technology (APT-CH) formulation, which uniquely combines FDA approved inactive ingredients with a designated active pharmaceutical ingredient to enhance efficacy against highly resistant microbes (both bacterial and fungal), it has proven effective in reducing fungal burden in skin tissue. This indicates its potential for reducing C. auris skin colonization (11).

Despite the in vitro evidence indicating the susceptibility of the C. auris to chlorhexidine (CHX), established guidelines for successful C. auris decolonization remain conspicuously absent, particularly within clinical settings, where patients undergoing daily CHX bathing continue to exhibit C. auris isolation from their skin, highlighting a notable discrepancy between in vitro findings and clinical outcomes (12).

The purpose of conducting this systematic review is to clarify to healthcare providers scientifically-sound information on the effectiveness of using chlorhexidine bathing in reducing the colonization of C. auris among affected patients in healthcare settings. By pooling the results of multiple studies and analysing the overall effect size of the intervention, a systematic review and meta-analysis can help synthesize the available evidence on the effectiveness of chlorhexidine bathing for decolonization of C. auris patients and instruct healthcare providers accordingly.

### Objectives

The primary aim of this study is to conduct a comprehensive systematic review and, if feasible, a meta-analysis, focusing on the efficacy of daily chlorhexidine bathing as an intervention for C. auris decolonization in adult patients within healthcare settings. To accomplish this objective, the following specific objectives will be addressed:

1. Perform a systematic review of full-length research articles published in peer-reviewed English-language scientific journals that investigate the effectiveness of daily chlorhexidine bathing for decolonizing C. auris in adult patients within healthcare settings.
2. Examine the potential impact of daily chlorhexidine bathing on reducing C. auris colonization, infection rates, and outbreaks among adult patients in healthcare settings.
3. Assess the degree to which daily chlorhexidine bathing contributes to the decolonization of C. auris among adult patients within healthcare settings, considering variations in the applied protocols.

Additionally, as part of the comprehensive evaluation of safety and potential risk factors, the following secondary objectives will be investigated:

1. Identify any adverse events associated with the implementation of daily chlorhexidine bathing as an intervention for C. auris decolonization among adult patients in healthcare settings.
2. Investigate the influence of potential risk factors, if identified, on the study outcomes by quantifying their effect sizes.

Furthermore, If the available data permit, we will further pursue the following objectives through the execution of a meta-analysis:

1. Calculate pooled effect sizes, such as risk ratios, odds ratios, or mean differences, to quantitatively assess the overall effectiveness of daily chlorhexidine bathing for Candida auris decolonization.
2. Conduct subgroup analyses to explore potential variations in effectiveness based on specific population characteristics (e.g., age, comorbidities), differences in chlorhexidine bathing protocols (e.g., concentration, frequency, duration), and geographical locations of the included studies.

## Methods and Analysis

This protocol adheres to the guidelines outlined in the Preferred Reporting Items for Systematic Reviews and Meta-analyses Protocol (PRISMA-P) (Supplemental file 1) (13). The study is scheduled to commence on November 1, 2023, and conclude on October 31, 2024. The final review process will be guided by the Cochrane Handbook for Systematic Reviews of Interventions and will align with the updated PRISMA 2020 statement (13,14).

### Eligibility criteria

A Population, Intervention, Comparator, and Outcomes (PICO) statement has been formulated to systematically assess the efficacy of chlorhexidine bathing in decreasing C. auris colonization among patients in healthcare settings who are affected by this pathogen.

#### Population

##### Inclusion

Studies involving adult patients (aged >18 years) admitted to healthcare facilities, including hospitals and long-term care facilities, who have been colonized with C. auris and treated with chlorhexidine bathing for C. auris decolonization.

##### Exclusion

To minimize potential confounding effects related to age, studies exclusively focusing on paediatric patients will be excluded.

#### Intervention

##### Inclusion

Studies employing chlorhexidine bathing in decolonizing C. auris among adult patients in healthcare setting.

##### Exclusion

Studies exploring decolonization methods other than chlorhexidine bathing or those failing to provide data on the effectiveness of chlorhexidine in C. auris decolonization will not be considered in this review.

#### Comparator

The comparator in this analysis will be standard hygiene practices or alternative decolonization methods other than chlorhexidine bathing, which may include routine bathing without chlorhexidine or the use of different topical agents for decolonization.

#### Outcome

##### Main Outcome

The primary outcome for this analysis will be the proportion of adult patients colonized with Candida auris who achieved successful decolonization when subjected to chlorhexidine bathing, quantified by effect size measures, as indicated by two consecutive negative test results with a one-week gap.

##### Additional Outcomes

1. The rate of adverse events or complications associated with chlorhexidine bathing among adult patients colonized with Candida auris.
2. Assess the effect of chlorhexidine bathing on Candida auris colonization rates, as well as its impact on the incidence of Candida auris infections and outbreaks among adult patients in healthcare settings.
3. Evaluate factors, both positively and negatively associated, with the efficacy of chlorhexidine bathing in C. auris decolonization, quantified through effect sizes such as risk ratios, odds ratios, or mean differences where applicable.

#### Types of studies

The studies should be published in English language and can include randomized controlled trials and non-randomized controlled trials. In contrast, studies published in non-English languages and those lacking peer review will be excluded. Furthermore, certain publication types, including reviews, editorials, letters, conference abstracts, case reports, case series, observational studies like cohort and case-control studies, and *in vivo* animal and *in vitro* studies will not be considered.

#### Information sources

We will conduct systematic searches in four electronic databases, including PubMed (National Library of Medicine), EMBASE (Elsevier), Scopus (Elsevier), Web of Science-Core Collection (Clarivate) to identify relevant studies that have been published in peer-reviewed scientific journals.

#### Search strategy

In January 2024, a comprehensive and systematic literature search will be carried out by MSS. Preliminary search term combinations, aligned with the PICO statement and the specified inclusion and exclusion criteria, have been identified and tested (Supplemental file 2). Preliminary searches were conducted on PubMed from June to August 2023, utilizing PubMed’s MeSH terms to ensure a systematic inclusion of relevant search terms and their synonyms. The final selection of search terms was reviewed by subject expert (BA). The search string developed for PubMed will be subsequently applied to all chosen databases, without applying geographical or publication year restrictions. To maximize retrieval, a combination of search fields, including ‘Title/Abstract’ and MeSH (or equivalent subject headings), will be employed. In the absence of MeSH, Thesaurus, or Subject Headings, a search field covering ‘Title’, ‘Abstract’, and ‘Keywords’ will be utilized. Reproducible search strings, along with results and notes for all databases included in the review, will be attached to the final study. To expand the search with a backward snowballing approach, the reference lists of included publications and identified systematic reviews will be manually searched. Prior to the final analysis and manual screening of the reference lists of included studies, a complete search update will be performed across all databases and will be presented in a PRISMA flow diagram.

## Study records

### Data management

The search results’ citations will be imported into the Covidence systematic review software, which will be used for record management, blinded screening, and conflict resolution. Covidence’s features, including automatic deduplication, title/abstract and full-text screening, and blinded conflict resolution, will facilitate our review process (15).

### Selection process

After the automatic removal of duplicate studies using Covidence, the unique studies retrieved will undergo a two-stage screening process conducted by two independent reviewers, MB and MSS, in accordance with the predetermined inclusion and exclusion criteria. The first stage will involve screening the titles and abstracts of the publications. In the second stage, the full texts of publications selected in the first stage will be assessed for inclusion, by the same independent reviewers. Any reasons for exclusion will be documented in the PRISMA flow diagram, as illustrated in Supplemental file 3. This diagram follows the guidelines outlined in The PRISMA 2020 statement (14). If discrepancies arise between the assessments of the two reviewers regarding study eligibility or reasons for exclusion, a third reviewer, BA, will resolve these discrepancies within the Covidence software. It’s important to note that the screening and conflict resolution modules in Covidence will be conducted in a blinded manner. Manual screening of the reference lists of all eligible reviewed articles, as well as of systematic reviews identified by the search, will be performed to find additional papers that meet the inclusion criteria.

### Data collection process

Two independent reviewers, MB and MSS, will autonomously extract data using a data extraction template created in Excel specifically for this study (Supplemental file 4). Prior to its implementation, this template will undergo a pilot test. Any discrepancies in the extracted data will be addressed and the data’s accuracy resolved through discussions involving the participation of other reviewer (BA). This collaborative process will continue until a consensus is reached, and all parties are in agreement.

### Data items

As a minimum requirement, the data extraction process will encompass essential information such as publication details (title, primary author’s name, publication year, and DOI), study characteristics (study type, type of treatment applied, data collection timeframe, measurement methods and outcomes, statistical analyses conducted, and any adjustments made for confounding variables), demographic characteristics of the study population, outcomes related to the application of daily chlorhexidine bathing for C. auris decolonization among adult patients within healthcare settings, the overall effect size of this intervention, any observed variations, and any declarations of conflict of interest or funding sources.

### Risk of bias in individual studies

Two independent reviewers, MB and MSS, will conduct a thorough evaluation of the risk of bias present in the studies included in this review. This assessment will be carried out employing the Cochrane risk-of-bias tool (16), a globally recognized instrument renowned for its meticulous and comprehensive approach to evaluating the methodological soundness of various study designs, through its four domains (Supplemental file 5). The tool’s well-structured framework facilitates the assessment of critical domains such as randomization, allocation concealment, blinding, management of incomplete outcome data, selective reporting, and the identification of other potential sources of bias. In the event of differences in risk of bias assessment between the two reviewers for specific studies, a third reviewer, BA, will act as an arbitrator following a comprehensive discussion. Simultaneously, the quality of evidence will undergo evaluation using the Grading of Recommendations Assessment, Development, and Evaluation (GRADE) scoring method, a systematic approach that involves all reviewers. This systematic assessment will ultimately enable the formulation of evidence-based summary statements.

### Pilot test

The study selection, data extraction, and assessment of RoB processes were tested using a 10% random sample (n=139) of studies identified through an initial pre-search covering all electronic databases. The results of this preliminary database search were uploaded to Covidence for further manual screening within the pilot sample. During the initial title/abstract screening in Covidence, the majority (n=68) of studies in the pilot sample were excluded by two reviewers (MB and MS), leaving 71 studies eligible for full-text screening after conflicts were resolved by a third reviewer (BA). Subsequently, during the full-text screening, 52 studies were excluded based on predefined exclusion criteria. Data extraction was carried out independently by two reviewers (MB and MS) for the remaining studies (n=4) that met the inclusion criteria outlined in the review protocol. The completed data extraction sheets were reviewed by the third reviewer (BA), and any discrepancies were resolved through discussions among the reviewers (MB, MSS, and BA) until a consensus was reached. Following this, two reviewers independently assessed the RoB of the eligible studies using the 4 domains outlined in the Cochrane RoB tool. Each domain in each of the eligible studies was assigned a RoB grade (low, probably low, probably high, or high). In cases where discrepancies arose between the judgments of the two reviewers, a third reviewer (BA) resolved them and also summarized the results of the RoB assessment. The results of this pilot study, including the study selection process, data extraction, and RoB assessment, have been provided in Supplementary Material 3, 4, and 5, respectively.

### Data synthesis

The eligible studies that meet our criteria will undergo a comprehensive narrative synthesis. This synthesis will involve the creation of summary tables that effectively communicate the key findings regarding the effectiveness of daily chlorhexidine bathing in decolonizing C. auris among adult patients in healthcare settings. These tables will encompass essential information related to the characteristics of the study population, treatment characteristics, outcomes pertaining to decolonization post-treatment, and other relevant findings.

In addition, if the data permit, we will conduct a meta-analysis to further examine the efficacy of chlorhexidine bathing for Candida auris decolonization, as well as the effect of potential risk factors, by estimating pooled effect sizes. When we identify two or more studies that provide suitable outcome estimates, two independent reviewers will rigorously assess their clinical comparability and, if appropriate, combine them for meta-analysis employing the inverse variance method and a random-effects model to account for any inter-study variations.

To gauge the degree of statistical heterogeneity among the included studies, we will employ the I^2^ statistics. The entire meta-analysis process will be executed using the RevMan software, and the synthesized results will be visually presented through forest plots.

Furthermore, as part of our analysis, we will employ a funnel plot to visually assess the presence of publication bias. In situations where outliers or asymmetry in the funnel plot are detected, we will conduct sensitivity analysis to explore the impact of these factors on our results.

### Analysis of subgroups or subsets

If the evidence suggests variations in the extent of decolonization based on specific population characteristics or variations in treatment protocols, this study will conduct sub-group analyses to delve deeper into these factors and their potential impact on decolonization outcomes. Additionally, sensitivity analyses will be performed to rigorously assess the robustness of our findings and ensure that they are not unduly influenced by specific factors or outliers. These analytical approaches will enhance the comprehensiveness and reliability of our study results.

### Quality of cumulative evidence

To ensure a comprehensive evaluation of the quality of evidence across all included studies, a team of at least six reviewers (MB, MSS, BA) will employ the Grading of Recommendations Assessment, Development, and Evaluation (GRADE) scoring method. This systematic approach will facilitate the generation of evidence-based summary statements, enhancing the overall rigor and reliability of our study’s conclusions.

### Patient and public involvement

This study will not involve the participation of patients or members of the public.

## ETHICS AND DISSEMINATION

Ethics approval is not required for this systematic review, given that it does not involve the participation of patients or the public. The study’s results will be disseminated through publication in a peer-reviewed journal and made available electronically. Furthermore, if the findings indicate a need for changes in the practice of daily chlorhexidine bathing for C. auris decolonization among adult patients in healthcare settings, a summary report will be shared with key healthcare and policy stakeholders in the United Arab Emirates.

## Strengths and limitations of this study

This study has several strengths:

1. Employing a rigorous and systematic search strategy, and strict adherence to established guidelines like PRISMA and GRADE, ensuring methodological rigor.
2. Engagement of multiple independent reviewers in screening, data extraction, bias assessment, and quality appraisal, enhancing result reliability.
3. Utilization of both narrative synthesis and meta-analysis for a comprehensive evidence examination.
4. Addressing a clinically vital matter by evaluating chlorhexidine bathing’s efficacy against C. auris colonization, with potential policy implications.

Nevertheless, the study does have limitations:

1. Potential publication bias due to reliance on peer-reviewed English-language articles, potentially missing relevant research.
2. Inherent variability in study populations, interventions, and outcomes across included studies may impact meta-analysis feasibility and limit the generalizability of the findings.
3. Findings are contingent on data availability and reporting within included studies, potentially limiting specific analyses and definitive conclusions.

## Data Availability

All data produced in the present study are available upon reasonable request to the authors

## Contributors

MB, MSS, and BA, actively participated in the conceptualization of the study, development of the research protocol, and the subsequent manuscript writing. They contributed to the design of the search strategy, conducted the literature search, performed screening of studies, and extracted data. In cases of conflicts, BA played a pivotal role in resolution.

## Patient consent for publication

Not applicable.

## Provenance and peer review

Not commissioned, externally peer reviewed.

## Supplemental material

## References

1. Cristina ML, Spagnolo AM, Sartini M, Carbone A, Oliva M, Schinca E, et al. An Overview on Candida auris in Healthcare Settings. Journal of Fungi [Internet]. 2023 Sep 8 [cited 2023 Oct 1];9(9):913. Available from: /pmc/articles/PMC10532978/

2. Chen J, Tian S, Han X, Chu Y, Wang Q, Zhou B, et al. Is the superbug fungus really so scary? A systematic review and meta-analysis of global epidemiology and mortality of Candida auris. BMC Infect Dis [Internet]. 2020 Dec 1 [cited 2023 Oct 2];20(1):1–10. Available from: https://bmcinfectdis.biomedcentral.com/articles/10.1186/s12879-020-05543-0

3. Rossow J, Ostrowsky B, Adams E, Greenko J, Mcdonald R, Vallabhaneni S, et al. Factors Associated With Candida auris Colonization and Transmission in Skilled Nursing Facilities With Ventilator Units, New York, 2016-2018. Clin Infect Dis [Internet]. 2021 Jun 1 [cited 2023 Oct 2];72(11):E753–60. Available from: https://pubmed.ncbi.nlm.nih.gov/32984882/

4. Southwick K, Adams EH, Greenko J, Ostrowsky B, Fernandez R, Patel R, et al. 2039. New York State 2016–2018: Progression from Candida auris Colonization to Bloodstream Infection. Open Forum Infect Dis [Internet]. 2018 Nov 26 [cited 2023 Oct 2];5(Suppl 1):S594. Available from: /pmc/articles/PMC6252412/

5. Information for Infection Preventionists | Fact Sheets | Candida auris | Fungal Diseases | CDC [Internet]. [cited 2023 Oct 15]. Available from: https://www.cdc.gov/fungal/candida-auris/fact-sheets/cdc-message-infection-experts.html

6. Du H, Bing J, Hu T, Ennis CL, Nobile CJ, Huang G. Candida auris: Epidemiology, biology, antifungal resistance, and virulence. PLoS Pathog [Internet]. 2020 Oct 22 [cited 2023 Oct 1];16(10). Available from: https://pubmed.ncbi.nlm.nih.gov/33091071/

7. Thoma R, Seneghini M, Seiffert SN, Vuichard Gysin D, Scanferla G, Haller S, et al. The challenge of preventing and containing outbreaks of multidrug-resistant organisms and Candida auris during the coronavirus disease 2019 pandemic: report of a carbapenem-resistant Acinetobacter baumannii outbreak and a systematic review of the literature. Antimicrob Resist Infect Control [Internet]. 2022 Dec 1 [cited 2023 Oct 1];11(1). Available from: https://pubmed.ncbi.nlm.nih.gov/35063032/

8. Ahmad S, Alfouzan W. Candida auris: Epidemiology, Diagnosis, Pathogenesis, Antifungal Susceptibility, and Infection Control Measures to Combat the Spread of Infections in Healthcare Facilities. Microorganisms [Internet]. 2021 [cited 2023 Oct 4];9(4). Available from: https://pubmed.ncbi.nlm.nih.gov/33920482/

9. Alp Ş, Arikan AkdaCli S. [Candida auris and Mechanisms of Antifungal Drug Resistance]. Mikrobiyol Bul [Internet]. 2021 [cited 2023 Oct 1];55(1):99–112. Available from: https://pubmed.ncbi.nlm.nih.gov/33590985/

10. Treatment and Management of C. auris Infections and Colonization | Candida auris | Fungal Diseases | CDC [Internet]. [cited 2023 Oct 1]. Available from: https://www.cdc.gov/fungal/candida-auris/c-auris-treatment.html

11. Elshaer M, Herrada J, Gamal A, McCormick TS, Ghannoum M. Efficacy of chlorhexidine in advanced performance technology formulation in decolonizing the skin using Candida auris skin colonization mouse model. Am J Infect Control [Internet]. 2023 Jul 1 [cited 2023 Oct 1];51(7):836–7. Available from: https://pubmed.ncbi.nlm.nih.gov/36417953/

12. Kean R, McKloud E, Townsend EM, Sherry L, Delaney C, Jones BL, et al. The comparative efficacy of antiseptics against Candida auris biofilms. Int J Antimicrob Agents [Internet]. 2018 Nov 1 [cited 2023 Oct 1];52(5):673–7. Available from: https://pubmed.ncbi.nlm.nih.gov/29775686/

13. Moher D, Shamseer L, Clarke M, Ghersi D, Liberati A, Petticrew M, et al. Preferred reporting items for systematic review and meta-analysis protocols (PRISMA-P) 2015 statement. Syst Rev [Internet]. 2015 [cited 2023 Oct 1];4(1):148–60. Available from: https://pubmed.ncbi.nlm.nih.gov/25554246/

14. Page MJ, McKenzie JE, Bossuyt PM, Boutron I, Hoffmann TC, Mulrow CD, et al. The PRISMA 2020 statement: an updated guideline for reporting systematic reviews. BMJ [Internet]. 2021 Mar 29 [cited 2023 Oct 1];372. Available from: https://www.bmj.com/content/372/bmj.n71

15. Covidence - Literature review management [Internet]. [cited 2023 Oct 1]. Available from: https://get.covidence.org/literature-review?campaignid=18238395256&adgroupid=138114520982&gclid=Cj0KCQjwjt-oBhDKARIsABVRB0yHy98EKShq75564p-7MysTlEVw8iZ0nTegIDhazEs55P6PHuOT2JUaAgeUEALw_wcB

16. Sterne JAC, Savović J, Page MJ, Elbers RG, Blencowe NS, Boutron I, et al. RoB 2: a revised tool for assessing risk of bias in randomised trials. BMJ [Internet]. 2019 [cited 2023 Oct 2];366. Available from: https://pubmed.ncbi.nlm.nih.gov/31462531/

