## Supplemental table 1 for "Chlorhexidine Bathing for Candida auris Decolonization among Adult Patients in Healthcare Settings: Protocol for a Systematic Review and Meta-Analysis": Candida Search String.docx

| **Database** | **Search string** | **Date, and number of hits** |
| --- | --- | --- |
| Pubmed | (“CHG-treat*”[Title/Abstract] OR “CHG treat*”[Title/Abstract] OR “CHG therap*”[Title/Abstract] OR “CHG-therap*”[Title/Abstract] OR “Chlorhexidin*”[Title/Abstract] OR “Chlorohexidin*”[Title/Abstract] OR “Chlorhexadin*”[Title/Abstract] OR “Chlorohexadin*”[Title/Abstract] OR “decolonis*”[Title/Abstract] OR “decoloniz*”[Title/Abstract] OR “bath*”[Title/Abstract] OR “Basin*”[Title/Abstract] OR “Balneotherap*”[Title/Abstract] OR “Rotersept*”[Title/Abstract] OR “Tubulicid*”[Title/Abstract] OR “Fimeil*”[Title/Abstract] OR “Hexadol*”[Title/Abstract] OR “Soretol*”[Title/Abstract] OR “Sterilon*”[Title/Abstract] OR “Nolvasan*”[Title/Abstract] OR “Cloresidina*”[Title/Abstract] OR “Sterido*”[Title/Abstract] OR “Savlon*”[Title/Abstract] OR “Hibistat*”[Title/Abstract] OR “Peridex*”[Title/Abstract] OR “R4KO0DY52L*”[Title/Abstract] OR “Biguanide*”[Title/Abstract] OR “imidodicarbonimid*”[Title/Abstract] OR “Savloclens*”[Title/Abstract] OR “Oro-Clense*”[Title/Abstract] OR “CCRIS 9230*”[Title/Abstract] OR “Merfen*”[Title/Abstract] OR “Nolvasan*”[Title/Abstract] OR “Diacetate*”[Title/Abstract] OR “EINECS 200-238-7*”[Title/Abstract] OR “R4KO0DY52L*”[Title/Abstract] OR “2826432*”[Title/Abstract] OR “Hibisol*”[Title/Abstract] OR “Decanoylacetaldehyde*”[Title/Abstract] OR “Prestwick*”[Title/Abstract] OR “Hibidil*”[Title/Abstract] OR “Hibisol*”[Title/Abstract] OR “Hibitane*”[Title/Abstract] OR “Hibiscrub*”[Title/Abstract] OR “Savloclens*”[Title/Abstract] OR “BSPBio*”[Title/Abstract] OR “9552079*”[Title/Abstract] OR “chloroanilino*”[Title/Abstract] OR “Tox21*”[Title/Abstract] OR “Hexamethylenebis*”[Title/Abstract] OR “Tetraazatetradeca*”[Title/Abstract] OR “Chlorophenyl*”[Title/Abstract] OR “0051301*”[Title/Abstract] OR “200828*”[Title/Abstract] OR “imidodicarbonimidic*”[Title/Abstract] OR “carbamimidamido*”[Title/Abstract] OR “methanimidamide*”[Title/Abstract] OR “Sebidin*”[Title/Abstract] OR “MK412A*”[Title/Abstract] OR "Chlorhexidine"[MeSH Terms] OR "chlorhexidine zinc acetate drug combination"[Supplementary Concept] OR "chlorhexidine phosphanilate"[Supplementary Concept] OR "chlorhexidine gluconate"[Supplementary Concept] OR "chlorhexidine gluconate lidocaine drug combination"[Supplementary Concept] OR "benzocaine chlorhexidine enoxolone drug combination"[Supplementary Concept] OR "chlorhexidine thymol drug combination"[Supplementary Concept] OR "Baths"[Mesh]) AND (“Candida-auris”[Title/Abstract] OR “Candida auris”[Title/Abstract] OR “C. auris”[Title/Abstract] OR “C.auris”[Title/Abstract] OR “C-auris”[Title/Abstract] OR “C auris”[Title/Abstract] OR "Candida auris infection" [Supplementary Concept] OR "Candida auris"[Mesh]) | 26 hits in 29/09/2023 |
| Scopus | ( TITLE-ABS-KEY ( "chg-treat*" OR "chg treat*" OR "chg therap*" OR "chg-therap*" OR "chlorhexidin*" OR "chlorohexidin*" OR "chlorhexadin*" OR "chlorohexadin*" OR "decolonis*" OR "decoloniz*" OR "bath*" OR "basin bed*" OR "balneotherap*" OR "c22h30cl2n10*" OR "rotersept*" OR "tubulicid*" OR "fimeil*" OR "hexadol*" OR "soretol*" OR "sterilon*" OR "nolvasan*" OR "cloresidina*" OR "sterido*" OR "merfen-incolore*" OR "savlon babycare*" OR "hibistat*" OR "dentisept*" OR "peridex*" OR "chembl790*" OR "r4ko0dy52l*" OR "mls001332388*" OR "chebi:3614*" OR "dtxsid203331*" OR "biguanide*" OR "ncgc00016246*" OR "smr000857146*" OR "imidodicarbonimid*" OR "dtxcid0013314*" OR "savloclens*" OR "superspray*" OR "hibispray*" OR "mls001304094*" OR "oro-clense*" OR "ccris 9230*" OR "houttuyfonamide*" OR "c22h30cl2n10*" OR "1246816-96-5*" OR "merfen-incolore*" OR "01000799135*" OR "nolvasan*" OR "diacetate*" OR "smr000718621*" OR "einecs 200-238-7*" OR "r4ko0dy52l*" OR "lisium*" OR "2826432*" OR "hibisol*" OR "dentisept*" OR "decanoylacetaldehyde*" OR "prestwick*" OR "hibidil*" OR "hibisol*" OR "hibitane*" OR "hibiscrub*" OR "hibispray*" OR "nsc526936*" OR "savloclens*" OR "schembl3984*" OR "bspbio*" OR "kbiogr*" OR "kbioss*" OR "mls001332387*" OR "mls002154209*" OR "divk1c_000761*" OR "spbio_000210*" OR "spbio_002185*" OR "bpbio1_000272*" OR "bdbm51937*" OR "bdbm64773*" OR "9552079*" OR "kbio1_000761*" OR "kbio2_000717*" OR "kbio2_003285*" OR "kbio2_005853*" OR "kbio3_001197*" OR "12303047*" OR "bdbm152706*" OR "hms1568m08*" OR "hms2095m08*" OR "hms2233b16*" OR "hms3712m08*" OR "chloroanilino*" OR "tox21*" OR "bdbm50170723*" OR "s5397*" OR "stk089248*" OR "hexamethylenebis*" OR "akos005394319*" OR "db00878*" OR "tetraazatetradeca*" OR "chlorophenyl*" OR "ncgc00016246*" OR "ncgc00091025*" OR "0051301*" OR "ab00053427*" OR "c06902*" OR "d07668*" OR "a830704*" OR "19626171*" OR "200828*" OR "01000799135*" OR "k52256627*" OR "01000799135*" OR "imidodicarbonimidic*" OR "carbamimidamido*" OR "methanimidamide*" OR "methanimidoyl*" ) AND TITLE-ABS-KEY ( "candida-auris" OR "candida auris" OR "c. auris" OR "c.auris" OR "c-auris" OR "c auris" OR "ca7lbn" OR "ca2lbn" OR "ca5lbn" OR "ca4lbn" OR "ca6lbn" OR "ca9lbn" OR "ca12lbn" OR "ca16lbn" OR " ca8lbn" OR "ca10lbn" OR "ca1lbn" OR "ca11lbn" OR "ca18lbn" OR "ca17lbn" OR "ca14lbn" OR "ca23lbn" OR "ca13lbn" OR "ca24lbn" OR "ca22lbn" OR "ca25lbn" OR "ca15lbn" OR "ca19lbn" OR "ca26lbn" OR "ca27lbn" OR "ca28lbn" OR "ca20lbn" OR "ca21lbn" OR "ca29lbn" OR "b13916" OR "b11205" OR "ca-am1" OR "b11220" OR "b12043" OR "b11809" OR "b13463" OR "b12037" OR "b12631" OR "b17721" OR "b11245" OR "b12342" ) ) | 56 hits in 29/09/2023 |
| Web of Science | “CHG-treat*” OR “CHG treat*” OR “CHG therap*” OR “CHG-therap*” OR “Chlorhexidin*” OR “Chlorohexidin*” OR “Chlorhexadin*” OR “Chlorohexadin*” OR “decolonis*” OR “decoloniz*” OR “bath*” OR “Basin Bed*” OR “Balneotherap*” OR “C22H30Cl2N10*” OR “Rotersept*” OR “Tubulicid*” OR “Fimeil*” OR “Hexadol*” OR “Soretol*” OR “Sterilon*” OR “Nolvasan*” OR “Cloresidina*” OR “Sterido*” OR “Merfen-incolore*” OR “Savlon babycare*” OR “Hibistat*” OR “Dentisept*” OR “Peridex*” OR “CHEMBL790*” OR “R4KO0DY52L*” OR “MLS001332388*” OR “CHEBI:3614*” OR “DTXSID203331*” OR “Biguanide*” OR “NCGC00016246*” OR “SMR000857146*” OR “imidodicarbonimid*” OR “DTXCID0013314*” OR “Savloclens*” OR “Superspray*” OR “Hibispray*” OR “MLS001304094*” OR “Oro-Clense*” OR “CCRIS 9230*” OR “Houttuyfonamide*” OR “C22H30Cl2N10*” OR “1246816-96-5*” OR “Merfen-incolore*” OR “01000799135*” OR “Nolvasan*” OR “Diacetate*” OR “SMR000718621*” OR “EINECS 200-238-7*” OR “R4KO0DY52L*” OR “Lisium*” OR “2826432*” OR “Hibisol*” OR “Dentisept*” OR “Decanoylacetaldehyde*” OR “Prestwick*” OR “Hibidil*” OR “Hibisol*” OR “Hibitane*” OR “Hibiscrub*” OR “Hibispray*” OR “NSC526936*” OR “Savloclens*” OR “SCHEMBL3984*” OR “BSPBio*” OR “KbioGR*” OR “KbioSS*” OR “MLS001332387*” OR “MLS002154209*” OR “DivK1c_000761*” OR “SPBio_000210*” OR “SPBio_002185*” OR “BPBio1_000272*” OR “BDBM51937*” OR “BDBM64773*” OR “9552079*” OR “KBio1_000761*” OR “KBio2_000717*” OR “KBio2_003285*” OR “KBio2_005853*” OR “KBio3_001197*” OR “12303047*” OR “BDBM152706*” OR “HMS1568M08*” OR “HMS2095M08*” OR “HMS2233B16*” OR “HMS3712M08*” OR “chloroanilino*” OR “Tox21*” OR “BDBM50170723*” OR “s5397*” OR “STK089248*” OR “Hexamethylenebis*” OR “AKOS005394319*” OR “DB00878*” OR “Tetraazatetradeca*” OR “Chlorophenyl*” OR “NCGC00016246*” OR “NCGC00091025*” OR “0051301*” OR “AB00053427*” OR “C06902*” OR “D07668*” OR “A830704*” OR “19626171*” OR “200828*” OR “01000799135*” OR “K52256627*” OR “01000799135*” OR “imidodicarbonimidic*” OR “carbamimidamido*” OR “methanimidamide*” OR “methanimidoyl*” (Topic) and “Candida-auris” OR “Candida auris” OR “C. auris” OR “C.auris” OR “C-auris” OR “C auris” OR “carlin” OR “carlin” OR “carlin” OR “carlin” OR “carlin” OR “ca9ln” OR “ca125bun” OR “CA16LBN” OR “ carlin” OR “ca10li” OR “carlin” OR “ca11sb9” OR “ca18li3” OR “CA17LBN” OR “ca14mn” OR “ca2clbn2” OR “ca3sbn” OR “ca2clbn2” OR “ca2clbn2” OR “ca2clbn2” OR “ca150b” OR “ca9ln” OR “ca2clbn2” OR “ca2clbn2” OR “ca2clbn2” OR “ca2clbn2” OR “ca2clbn2” OR “ca2clbn2” OR “b1916” OR “b11202” OR “CA-AM1” OR “b11202” OR “b1204d” OR “b11209” OR “b1343” OR “b1237” OR “b12e31” OR “bb7721” OR “b11244” OR “ba2342” (Topic) | 27 hits in 29/09/2023 |
| Embase | ('chg-treat*':ti,ab,kw OR 'chg treat*':ti,ab,kw OR 'chg therap*':ti,ab,kw OR 'chg-therap*':ti,ab,kw OR 'chlorhexidin*':ti,ab,kw OR 'chlorohexidin*':ti,ab,kw OR 'chlorhexadin*':ti,ab,kw OR 'chlorohexadin*':ti,ab,kw OR 'decolonis*':ti,ab,kw OR 'decoloniz*':ti,ab,kw OR 'bath*':ti,ab,kw OR 'basin bed*':ti,ab,kw OR 'balneotherap*':ti,ab,kw OR 'rotersept*':ti,ab,kw OR 'tubulicid*':ti,ab,kw OR 'fimeil*':ti,ab,kw OR 'hexadol*':ti,ab,kw OR 'soretol*':ti,ab,kw OR 'sterilon*':ti,ab,kw OR 'cloresidina*':ti,ab,kw OR 'sterido*':ti,ab,kw OR 'savlon babycare*':ti,ab,kw OR 'hibistat*':ti,ab,kw OR 'peridex*':ti,ab,kw OR 'chembl790*':ti,ab,kw OR 'mls001332388*':ti,ab,kw OR 'chebi:3614*':ti,ab,kw OR 'dtxsid203331*':ti,ab,kw OR 'biguanide*':ti,ab,kw OR 'smr000857146*':ti,ab,kw OR 'imidodicarbonimid*':ti,ab,kw OR 'dtxcid0013314*':ti,ab,kw OR 'superspray*':ti,ab,kw OR 'mls001304094*':ti,ab,kw OR 'oro-clense*':ti,ab,kw OR 'ccris 9230*':ti,ab,kw OR 'houttuyfonamide*':ti,ab,kw OR 'c22h30cl2n10*':ti,ab,kw OR '1246816-96-5*':ti,ab,kw OR 'merfen-incolore*':ti,ab,kw OR 'nolvasan*':ti,ab,kw OR 'diacetate*':ti,ab,kw OR 'smr000718621*':ti,ab,kw OR 'einecs 200-238-7*':ti,ab,kw OR 'r4ko0dy52l*':ti,ab,kw OR 'lisium*':ti,ab,kw OR '2826432*':ti,ab,kw OR 'dentisept*':ti,ab,kw OR 'decanoylacetaldehyde*':ti,ab,kw OR 'prestwick*':ti,ab,kw OR 'hibidil*':ti,ab,kw OR 'hibisol*':ti,ab,kw OR 'hibitane*':ti,ab,kw OR 'hibiscrub*':ti,ab,kw OR 'hibispray*':ti,ab,kw OR 'nsc526936*':ti,ab,kw OR 'savloclens*':ti,ab,kw OR 'schembl3984*':ti,ab,kw OR 'bspbio*':ti,ab,kw OR 'kbiogr*':ti,ab,kw OR 'kbioss*':ti,ab,kw OR 'mls001332387*':ti,ab,kw OR 'mls002154209*':ti,ab,kw OR 'divk1c_000761*':ti,ab,kw OR 'spbio_000210*':ti,ab,kw OR 'spbio_002185*':ti,ab,kw OR 'bpbio1_000272*':ti,ab,kw OR 'bdbm51937*':ti,ab,kw OR 'bdbm64773*':ti,ab,kw OR '9552079*':ti,ab,kw OR 'kbio1_000761*':ti,ab,kw OR 'kbio2_000717*':ti,ab,kw OR 'kbio2_003285*':ti,ab,kw OR 'kbio2_005853*':ti,ab,kw OR 'kbio3_001197*':ti,ab,kw OR '12303047*':ti,ab,kw OR 'bdbm152706*':ti,ab,kw OR 'hms1568m08*':ti,ab,kw OR 'hms2095m08*':ti,ab,kw OR 'hms2233b16*':ti,ab,kw OR 'hms3712m08*':ti,ab,kw OR 'chloroanilino*':ti,ab,kw OR 'tox21*':ti,ab,kw OR 'bdbm50170723*':ti,ab,kw OR 's5397*':ti,ab,kw OR 'stk089248*':ti,ab,kw OR 'hexamethylenebis*':ti,ab,kw OR 'akos005394319*':ti,ab,kw OR 'db00878*':ti,ab,kw OR 'tetraazatetradeca*':ti,ab,kw OR 'chlorophenyl*':ti,ab,kw OR 'ncgc00016246*':ti,ab,kw OR 'ncgc00091025*':ti,ab,kw OR '0051301*':ti,ab,kw OR 'ab00053427*':ti,ab,kw OR 'c06902*':ti,ab,kw OR 'd07668*':ti,ab,kw OR 'a830704*':ti,ab,kw OR '19626171*':ti,ab,kw OR '200828*':ti,ab,kw OR 'k52256627*':ti,ab,kw OR '01000799135*':ti,ab,kw OR 'imidodicarbonimidic*':ti,ab,kw OR 'carbamimidamido*':ti,ab,kw OR 'methanimidamide*':ti,ab,kw OR 'methanimidoyl*':ti,ab,kw) AND ('candida-auris':ti,ab,kw OR 'candida auris':ti,ab,kw OR 'c. auris':ti,ab,kw OR 'c.auris':ti,ab,kw OR 'c-auris':ti,ab,kw OR 'c auris':ti,ab,kw OR 'ca7lbn':ti,ab,kw OR 'ca2lbn':ti,ab,kw OR 'ca5lbn':ti,ab,kw OR 'ca4lbn':ti,ab,kw OR 'ca6lbn':ti,ab,kw OR 'ca9lbn':ti,ab,kw OR 'ca12lbn':ti,ab,kw OR 'ca16lbn':ti,ab,kw OR 'ca8lbn':ti,ab,kw OR 'ca10lbn':ti,ab,kw OR 'ca1lbn':ti,ab,kw OR 'ca11lbn':ti,ab,kw OR 'ca18lbn':ti,ab,kw OR 'ca17lbn':ti,ab,kw OR 'ca14lbn':ti,ab,kw OR 'ca23lbn':ti,ab,kw OR 'ca13lbn':ti,ab,kw OR 'ca24lbn':ti,ab,kw OR 'ca22lbn':ti,ab,kw OR 'ca25lbn':ti,ab,kw OR 'ca15lbn':ti,ab,kw OR 'ca19lbn':ti,ab,kw OR 'ca26lbn':ti,ab,kw OR 'ca27lbn':ti,ab,kw OR 'ca28lbn':ti,ab,kw OR 'ca20lbn':ti,ab,kw OR 'ca21lbn':ti,ab,kw OR 'ca29lbn':ti,ab,kw OR 'b13916':ti,ab,kw OR 'b11205':ti,ab,kw OR 'ca-am1':ti,ab,kw OR 'b11220':ti,ab,kw OR 'b12043':ti,ab,kw OR 'b11809':ti,ab,kw OR 'b13463':ti,ab,kw OR 'b12037':ti,ab,kw OR 'b12631':ti,ab,kw OR 'b17721':ti,ab,kw OR 'b11245':ti,ab,kw OR 'b12342':ti,ab,kw) | 38 hits in 29/09/2023 |

Initial Plan

**“CHG-treat*” OR “CHG treat*” OR “CHG therap*” OR “CHG-therap*” OR “Chlorhexidin*” OR “Chlorohexidin*” OR “Chlorhexadin*” OR “Chlorohexadin*” OR “decolonis*” OR “decoloniz*” OR “bath*” OR “Basin Bed*” OR “Balneotherap*” OR “C22H30Cl2N10*” OR “Rotersept*” OR “Tubulicid*” OR “Fimeil*” OR “Hexadol*” OR “Soretol*” OR “Sterilon*” OR “Nolvasan*” OR “Cloresidina*” OR “Sterido*” OR “Merfen-incolore*” OR “Savlon babycare*” OR “Hibistat*” OR “Dentisept*” OR “Peridex*” OR “CHEMBL790*” OR “R4KO0DY52L*” OR “MLS001332388*” OR “CHEBI:3614*” OR “DTXSID203331*” OR “Biguanide*” OR “NCGC00016246*” OR “SMR000857146*” OR “imidodicarbonimid*” OR “DTXCID0013314*” OR “Savloclens*” OR “Superspray*” OR “Hibispray*” OR “MLS001304094*” OR “Oro-Clense*” OR “CCRIS 9230*” OR “Houttuyfonamide*” OR “C22H30Cl2N10*” OR “1246816-96-5*” OR “Merfen-incolore*” OR “01000799135*” OR “Nolvasan*” OR “Diacetate*” OR “SMR000718621*” OR “EINECS 200-238-7*” OR “R4KO0DY52L*” OR “Lisium*” OR “2826432*” OR “Hibisol*” OR “Dentisept*” OR “Decanoylacetaldehyde*” OR “Prestwick*” OR “Hibidil*” OR “Hibisol*” OR “Hibitane*” OR “Hibiscrub*” OR “Hibispray*” OR “NSC526936*” OR “Savloclens*” OR “SCHEMBL3984*” OR “BSPBio*” OR “KbioGR*” OR “KbioSS*” OR “MLS001332387*” OR “MLS002154209*” OR “DivK1c_000761*” OR “SPBio_000210*” OR “SPBio_002185*” OR “BPBio1_000272*” OR “BDBM51937*” OR “BDBM64773*” OR “9552079*” OR “KBio1_000761*” OR “KBio2_000717*” OR “KBio2_003285*” OR “KBio2_005853*” OR “KBio3_001197*” OR “12303047*” OR “BDBM152706*” OR “HMS1568M08*” OR “HMS2095M08*” OR “HMS2233B16*” OR “HMS3712M08*” OR “chloroanilino*” OR “Tox21*” OR “BDBM50170723*” OR “s5397*” OR “STK089248*” OR “Hexamethylenebis*” OR “AKOS005394319*” OR “DB00878*” OR “Tetraazatetradeca*” OR “Chlorophenyl*” OR “NCGC00016246*” OR “NCGC00091025*” OR “0051301*” OR “AB00053427*” OR “C06902*” OR “D07668*” OR “A830704*” OR “19626171*” OR “200828*” OR “01000799135*” OR “K52256627*” OR “01000799135*” OR “imidodicarbonimidic*” OR “carbamimidamido*” OR “methanimidamide*” OR “methanimidoyl*”**

**And**

**“Candida-auris” OR “Candida auris” OR “C. auris” OR “C.auris” OR “C-auris” OR “C auris” OR “CA7LBN” OR “CA2LBN” OR “CA5LBN” OR “CA4LBN” OR “CA6LBN” OR “CA9LBN” OR “CA12LBN” OR “CA16LBN” OR “ CA8LBN” OR “CA10LBN” OR “CA1LBN” OR “CA11LBN” OR “CA18LBN” OR “CA17LBN” OR “CA14LBN” OR “CA23LBN” OR “CA13LBN” OR “CA24LBN” OR “CA22LBN” OR “CA25LBN” OR “CA15LBN” OR “CA19LBN” OR “CA26LBN” OR “CA27LBN” OR “CA28LBN” OR “CA20LBN” OR “CA21LBN” OR “CA29LBN” OR “B13916” OR “B11205” OR “CA-AM1” OR “B11220” OR “B12043” OR “B11809” OR “B13463” OR “B12037” OR “B12631” OR “B17721” OR “B11245” OR “B12342”**
